# Dim Light Melatonin Onset Profile Phenotypes in a Real-World Clinical Population with Home-Based, Self-Collected Salivary Assessments: Identification, Prevalence, and Potential Clinical Relevance

**DOI:** 10.64898/2025.12.19.25342700

**Authors:** Ellen R. Stothard, Chris S. Schwartz, Michele L. Okun, Steve W. Granger, Benjamin C. Wiegand, Yannan Liu, David E. McCarty, Robert J. Thomas

## Abstract

Recent methodological advances have enabled minimally invasive, convenient self-assessment of central circadian phase in home-based settings [1]. In this report, we describe salivary melatonin onset secretion profiles in 261 participants (36% male; age range, 9–83 years) who were recruited from 18 clinics in North America or by self-referral. All participants received a standardized, at-home central circadian phase assessment kit by mail or directly through their provider, along with written and video sample collection instructions.

The standard protocol consisted of 7 or 9 saliva samples collected at 1-hour intervals in dim light on a single occasion, starting up to 7 hours prior to an individual’s habitual bedtime and proceeding until 1–5 hours past bedtime. The number of samples and the start and end times were highly dependent on the nature and timing of the individual’s primary concern and were dictated by the referring sleep-health provider. Samples were frozen immediately after collection and then returned to a central laboratory via 2-day shipping after an additional 48 hours of home freezing. Samples were subsequently kept frozen until the day of melatonin assay.

Of the 261 participants, 91 (34.9%) exhibited DLMOs within the Predicted Onset Window (POW). The remaining 170 participants (65.2%) showed profiles that did not meet criteria for a predicted, aligned phase onset, including 162 (62.1% of the total sample) that could be classified into atypical phenotypes, distributed as follows: 28 showed a Delayed Melatonin Onset (10.7%), 28 showed an Advanced Melatonin Onset (10.7%), 27 exhibited Hypermelatoninemia (10.3%), 36 exhibited Hypomelatoninemia (13.8%), and 43 presented with Irregular/Multipeak profiles (16.5%). The remaining 8 participants (3.1%) exhibited profiles that could not be reliably classified.

These results highlight the unexpectedly high proportion of non-predicted melatonin patterns and demonstrate that over 80% of profiles can be reliably assigned to clinically meaningful circadian phenotypes. The Discussion explores how real-world melatonin profiling can identify relevant circadian contributors to sleep disruption that symptom reports alone may fail to detect. We also describe and discuss these phenotypes in detail, considering their biological contexts and potential clinical relevance.

## INTRODUCTION

The field of circadian medicine recognizes the central role of biological timing in the regulation of health and disease [2–4]. Dim Light Melatonin Onset (DLMO) is considered a gold standard for characterizing central circadian phase [1, 5–8]. DLMO reflects the evening rise in endogenous melatonin and provides a precise physiological signature for evaluating internal clock timing and the onset of the biological night. Accurate assessment of circadian phase is considered necessary for evaluating circadian rhythm sleep–wake disorders (CRSWDs), informing intervention strategies, and addressing misaligned sleep patterns [9]. The clinical utility of DLMO assessment is well validated in sleep medicine, where its implementation has traditionally required extended time in light-restricted environments, professional healthcare staff, and the collection of repeated venous blood specimens.

A critical barrier in this area is also linguistic and conceptual. In clinical environments, certain ideas, particularly those involving non-linear or circular time, remain difficult to conceptualize and, consequently, difficult to discuss. Because they are hard to articulate, such concepts are often avoided altogether. This phenomenon has been noted in medical education, where complexity is under-addressed precisely because of its difficulty of articulation [10]. Our work suggests that home-based salivary DLMO profiling provides not only biological data, but also a shared language and process through which clinicians and patients can begin to navigate these complex, circular-time phenomena and their associated biological implications. This approach makes it possible to move conversations about circadian misalignment out of the realm of inaccessible medical jargon and into pragmatic, collaborative clinical practice [11].

Multiple independent validation studies have demonstrated that research-grade, at-home saliva collection, when paired with patient education, structured protocols, lighting control, and centralized assay processing, can yield reliable DLMO estimates for circadian phase assessment [1, 8, 12–14]. To date, however, few studies have described the range of salivary melatonin profiles that emerge when DLMO assessments are applied in real-world, non-clinical settings. There are several reasons to consider that this variation may differ from that observed when DLMO is implemented as a laboratory-based procedure rather than as a home-based self-assessment. The aim of this study was to assess the feasibility of implementing this highly standardized procedure outside the clinic and to examine inter-individual variation in results.

Our literature review suggested that at least five mutually exclusive DLMO phenotype profiles exist [15–20]. Building on this, we utilize a framework in which the established physiological pattern of melatonin onset supports a practical classification system, then evaluated whether additional patterns emerged in a real-world clinical dataset. Specifically, we group participants into five theoretical phenotype profiles, estimate their relative prevalence within a broader clinical population, and aggregate salivary melatonin concentration ranges across individuals within each phenotype. This approach enables the development of a reference database that can inform future best practice standards.

## METHODS

### Overview

This study represents a secondary data analysis of CLIA-certified laboratory data accumulated during the pilot phase of a commercial DLMO assessment program provided by Salimetrics LLC (for more details, see biologyofsleep.com). The program was made available to physicians in North America in 2022 and enabled home-based saliva collection and centralized laboratory processing of samples for melatonin to evaluate circadian phase in patients with sleep disruption.

Participants undergoing DLMO assessment were enrolled in the study after agreeing to the approved informed consent form included with the saliva collection kit. This permitted their DLMO data to be retained in a master database for subsequent secondary analyses along with a brief questionnaire assessing demographic characteristics and medication use. Physicians and participants received laboratory results directly via a HIPAA-compliant portal. The project was approved by Western IRB (IRB Tracking # 20233850).

### Participants

There were 261 eligible participants included in the current analyses, randomly selected from a total pool (n = 324) of DLMO results issued between April 19, 2023, and July 9, 2024. Results issued prior to April 19, 2023, did not include a request to report medication or melatonin use and were therefore excluded from analysis.

Because patients present with individualized sleep–wake patterns, daily routines, and symptom profiles, the sample scheduling component of the protocol was designed to be tailored to each participant’s primary concern and habitual sleep timing. Anchoring sample timing to bedtime, rather than clock time, optimized circadian alignment across participants. When warranted, physicians could also select alternative protocols, such as a 24-hour Circadian Phase Map or Dim Light Melatonin Offset (DLMOff).

As a result, sample collection timing varied across participants. The inclusion criterion applied to the broader dataset was as follows: of the 324 eligible datasets, 261 were selected based on participants having completed a DLMO protocol in which 7 or 9 samples were collected within 7 hours before and extending up to 5 hours after the individual’s average habitual bedtime. This sampling window allowed for standardized evaluation of circadian phase relative to each participant’s typical sleep–wake schedule.

### Procedure

Each consenting participant received a standardized saliva collection kit by mail, accompanied by both written and video instructions (see Supplemental Material and biologyofsleep.com). The standard protocol established a collection window starting either 5 or 7 hours prior to the participant’s routine bedtime, during which either 7 or 9 saliva samples were collected at hourly intervals. The sampling start and end times were determined by the referring provider based on the individual’s primary concern and initial clinical evaluation.

Participants were instructed to follow their healthcare provider’s guidance regarding withdrawal from melatonin supplements, as well as other medications such as sleep aids, NSAIDs, sedative-hypnotic medications, β1-adrenergic blockers, and/or other relevant medications. In general, a washout period of 10 days was recommended for melatonin supplements and 3 days for other medications, when safe and comfortable to do so. Medication name, dosage, and timing on the day of sampling and during the 24 hours prior to sampling were recorded by the participant on the sampling form.

During the collection window, beverages were restricted to water only. Following Murray and colleagues [1, 12, 13], participants were instructed to avoid caffeinated beverages or foods, alcohol, pitted fruits, chocolate, and bananas within 8 hours prior to collecting the first sample and throughout the sampling window. If food was consumed during the assessment, participants were instructed to avoid large meals within 1 hour of sample collection and to consume any snack items immediately after sampling, followed by rinsing the mouth with water before the next sample collection. Participants were also directed to maintain ambient light levels below approximately 10 lux (i.e., about as bright as a small nightlight) during the collection window. Instructions specified that participants should close all blinds and curtains and dim lights to the lowest level sufficient to read instructions, and turn off other indoor lights (e.g., bathroom, kitchen). Screens such as televisions, computer monitors, and mobile devices were permitted, provided they were set to the lowest brightness setting. Sunglasses were recommended if screens were used, as an additional layer of protection from light exposure (see Supplemental Material for additional detail).

Participants were instructed to freeze samples immediately after each collection using a pre-frozen, insulated storage and transport container validated to maintain appropriate temperatures for 2-day transport without dry ice. Samples were shipped via 2-day express delivery to a centralized CLIA-certified laboratory (Salimetrics LLC, State College, PA), where they were thawed, centrifuged, and analyzed for melatonin concentration (pg/mL) using a high-sensitivity ELISA (Salimetrics catalog #1-3402; limit of detection, 1.37 pg/mL; intra-assay and inter-assay coefficients of variation <5.20% and <8.26%, respectively).

A printed sampling form and questionnaire were included with the assessment to capture demographic information, routine bedtime, sample collection times, and medication and supplement use.

### Melatonin Onset Classification

Following the consensus of Lewy, Pandi-Perumal, and colleagues [5, 7], we defined *melatonin onset* as the clock time at which melatonin levels rise above a calculated threshold representing an increase of two standard deviations above the mean of three daytime baseline samples (the *3k method*) [1, 5–7, 21]. When this criterion could not be applied, onset was defined using a fixed threshold of 4 pg/mL, particularly when baseline samples were insufficient or fell below the assay limit of detection.

We anticipated the following profile categories: **Predicted Onset Window DLMO (POW DLMO)** defined as melatonin onset occurring within 1–3 hours before habitual bedtime, with peak melatonin concentrations exceeding 10 pg/mL; **Delayed Melatonin Onset** defined as melatonin onset occurring within 1 hour of, at, or after self-reported bedtime; and **Advanced Melatonin Onset** defined as melatonin onset occurring more than 3 hours before reported bedtime. Similar patterns have been well documented in the DLMO clinical literature using traditional implementation methods in controlled laboratory environments [8, 16, 20].

In addition, we anticipated two phenotype profiles reflecting the potential influence of medications that either directly or indirectly (via iatrogenic effects) elevate or attenuate melatonin secretion [22, 23]. **Hypermelatoninemia** was defined as elevated melatonin levels (>10 pg/mL) without a clear onset pattern across more than 85% of samples, whereas **Hypomelatoninemia** was defined as little to no consistent rise in melatonin, or melatonin levels that remained consistently below 10 pg/mL for more than 85% of sampling time points. Importantly, this latter pattern may also be consistent with melatonin suppression resulting from extended bright-light exposure during the sample collection period.

Participants were categorized as POW DLMO, Delayed Melatonin Onset, or Advanced Melatonin Onset phenotypes based on the presence of a calculated DLMO. Remaining participants were assigned to the Hypermelatoninemia or Hypomelatoninemia groups based on whether melatonin concentrations were predominantly above or below 10 pg/mL at more than 85% of the 7 or 9 sampled time points.

Classification was performed by the Salimetrics sleep-health program director, who was blinded to participant identity and clinical or demographic data. To assess classification reliability, 52 profiles (at least eight profiles from each phenotype group, except “Other”) were randomly selected and independently evaluated by a second blinded rater using the same decision rules. Interrater agreement was 92.3% across categories. Discrepant classifications (N = 2) within the reliability subsample were resolved by consensus discussion.

An additional phenotype was defined post hoc based on content analysis: **Irregular/Multipeak**, characterized by unstable melatonin profiles lacking a clear onset and exhibiting multiple small peaks or erratic fluctuations in melatonin levels.

A small subset of profiles was categorized as **“Other/Unclassified”** if they demonstrated a gradual, steadily increasing trend across the full sampling window without a clear physiological inflection point or a statistically distinguishable rise above baseline. Interrater agreement for this post hoc category was 80%. The “Other” classification accounted for 8 of the 261 total profiles, as these cases did not meet criteria for assignment to any of the predefined phenotypes.

## RESULTS

The resulting sample (N = 261) was 36% male and ranged in age from 9 to 83 years (median age = 47.4 years; see Table 1). Of the 261 participants, 91 profiles exhibited a DLMO occurring within the Predicted Onset Window (POW; 34.8%). The remaining 62.1% (N = 162) presented with melatonin onset patterns that occurred outside the predicted onset window or exhibited no observable onset, including 28 with Delayed Melatonin Onset (10.7%), 28 with Advanced Melatonin Onset (10.7%), 27 with Hypermelatoninemia (10.3%), 36 with Hypomelatoninemia (13.8%), 43 with Irregular or Multipeak Patterns (16.5%), and 8 profiles (3.1%) that could not be classified.

**Table 1.** Age Distribution of the Study Participants.

| **Age** | **# of Profiles** | **% of Total** |
| --- | --- | --- |
| 0--17 | 6 | 2.30% |
| 18--29 | 46 | 17.62% |
| 30--44 | 65 | 24.90% |
| 45--59 | 69 | 26.44% |
| 60--74 | 61 | 23.37% |
| 75+ | 13 | 4.98% |
| Age Not Provided | 1 | 0.38% |

**Table 2.** Profile Distribution by POW DLMO and Atypical Phenotypes.

| Phenotype | # of Profiles |
| --- | --- |
| POW DLMO | 91 |
| Atypical | 162 |
| Delayed Melatonin Onset | 28 |
| Advanced Melatonin Onset | 28 |
| Hypermelatoninemia | 27 |
| Hypomelatoninemia | 36 |
| Irregular/Multipeak | 43 |
| Other/Unclassified | 8 |

Within the Hypermelatoninemia group, 96.3% of participants reported taking medications that may influence circulating melatonin levels, including 63% who were taking psychiatric medications and 37% who had taken melatonin supplements within the past 10 days. Similarly, 92.7% of participants in the Hypomelatoninemia group reported use of medications with potential influence on melatonin levels, including 58.3% who were taking psychiatric medications.

## DISCUSSION

To our knowledge, this is among the first studies to characterize salivary melatonin onset phenotypes in a mixed clinical and general population under real-world sampling conditions. Our analysis suggests that in 96.9% of cases, inter-individual variability in melatonin onset could be reliably classified into distinct patterns using decision rules based on DLMO timing, melatonin concentration levels, and degree of inter-sample change. This represents an opportunity for providers and digital health tools to deliver objective phenotypic classifications that may guide more efficient and targeted treatment.

Notably, the majority of participants (approximately two-thirds) did not exhibit a melatonin onset pattern within the predicted onset window: roughly one in five demonstrated an early or delayed shift; one in four exhibited hyper- or hypomelatoninemic profiles without a clear onset; and one in six showed variable melatonin levels with no clear peak. Collectively, these phenotypes illustrate how real-world melatonin profiling can identify relevant circadian contributors to sleep disruption that symptom reports alone often fail to reveal. Below, we describe and discuss these phenotypes in detail, considering their biological contexts and potential clinical relevance.

Ninety-one participants exhibited a melatonin onset occurring within 1–3 hours prior to habitual bedtime, with peak concentrations exceeding 10 pg/mL (see Figure 1). This Predicted Onset Window (POW) DLMO profile suggests physiologically typical circadian timing and amplitude and was observed across a broad age range in both males and females. Importantly, the presence of predicted melatonin rhythms in individuals seeking care for sleep-related complaints suggests that circadian misalignment is not the sole driver of their sleep disruption. Instead, these cases highlight the potential contribution of behavioral, psychological, medical, or environmental factors that may impair sleep despite intact circadian regulation. Identifying circadian profiles in symptomatic individuals underscores the value of DLMO testing for distinguishing circadian-driven sleep disturbances from those rooted in other mechanisms.

**Figure 1.**
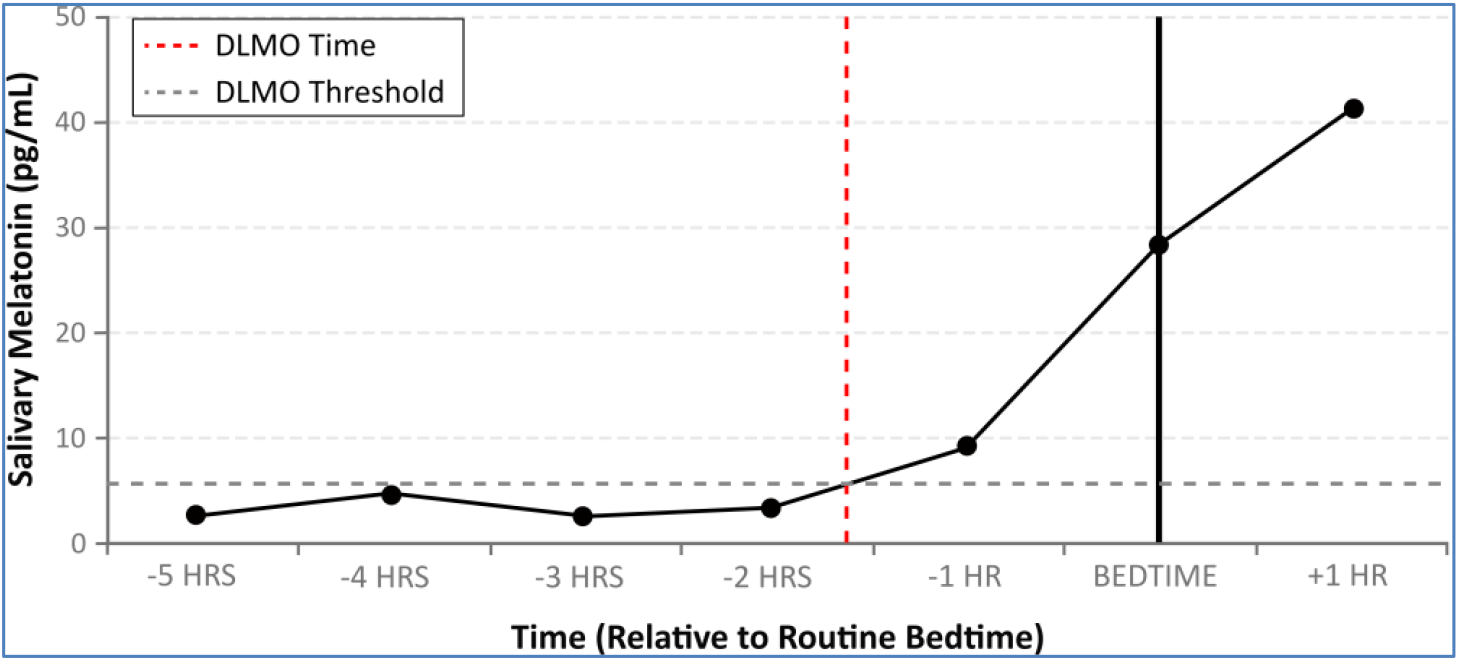
Predicted Onset Window DLMO Profile. Representative profile within the Predicted Onset Window group for melatonin profiles where DLMO is calculated to be within 1-3 hours prior to bedtime. Vertical dashed line represents DLMO time, and horizontal dashed line represents either the calculated or cutoff threshold.

Twenty-eight participants exhibited a Delayed Melatonin Onset (see Figure 2), defined by melatonin onset occurring within 1 hour of, at, or after their average reported bedtime, indicating a misalignment in which the biological night lags behind behavioral expectations. This delay may contribute to prolonged sleep latency, difficulty with morning awakenings, and associated daytime impairment—symptoms often misattributed to insomnia or poor sleep hygiene. Delayed DLMO profiles align with features of Delayed Sleep–Wake Phase Disorder (DSWPD) and may serve as an objective biomarker for identifying individuals at risk for circadian-related sleep difficulties. However, for future clinical implementation, it will be important to confirm adherence to dim-light conditions during early evening collection, as light exposure prior to sunset may suppress melatonin onset. The presence of delayed phase profiles in this sample underscores the importance of considering circadian delay as a distinct contributor to sleep complaints, particularly when sleep initiation difficulties might otherwise be attributed to behavioral or psychological factors.

**Figure 2.**
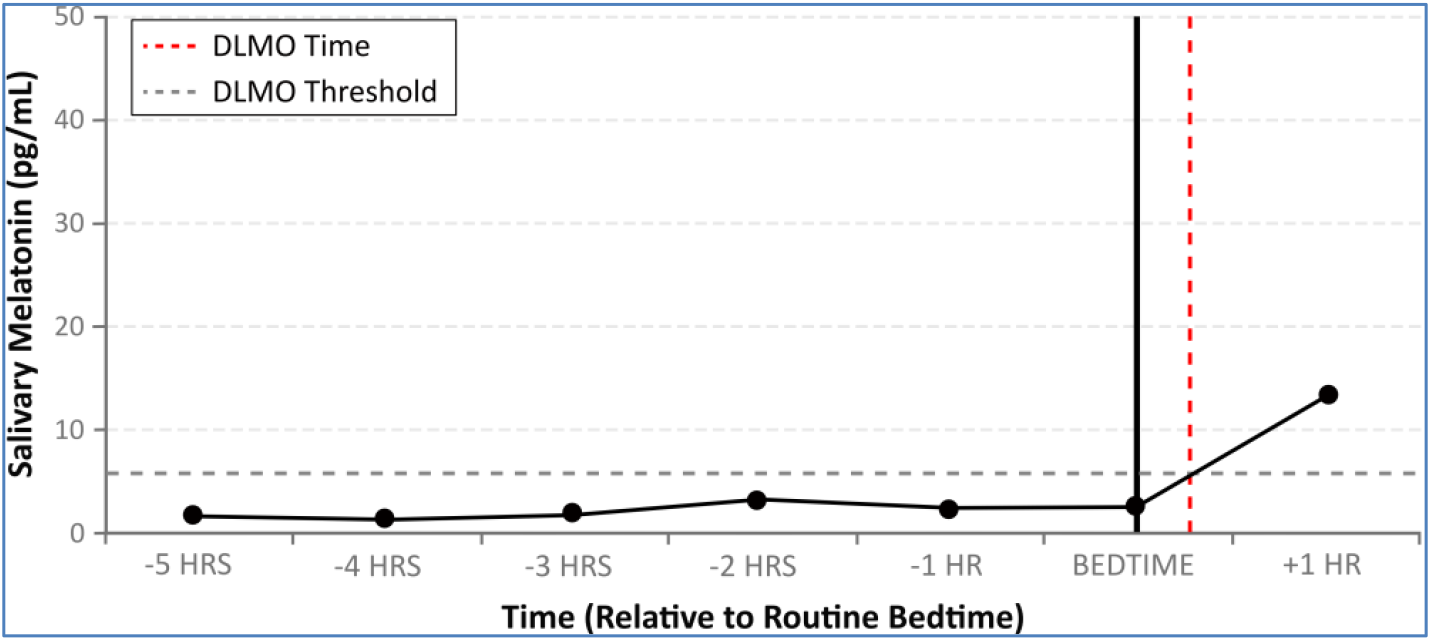
Delayed Melatonin Onset Profile. Representative profile within the Delayed Melatonin Onset group for melatonin profiles where DLMO is calculated to be within 1 hour prior to bedtime, at bedtime, or 1 hour after bedtime. Vertical dashed line represents DLMO time and horizontal dashed line represents either the calculated or cutoff threshold.

Twenty-eight participants also exhibited an Advanced Melatonin Onset profile (see Figure 3), characterized by a melatonin rise occurring more than 3 hours before reported bedtime. Such misalignment between biological night and behavioral sleep timing may contribute to early evening sleepiness, early morning awakenings, and daytime fatigue—symptoms often misattributed to poor sleep quality, aging, or insomnia. While these profiles are consistent with an advanced sleep phase phenotype, a substantial proportion of individuals, nearly half in some clinical settings [4, 24, 25], may instead present clinically as phase delayed, owing to difficulty maintaining circadian entrainment and adherence to a sleep schedule that conflicts with endogenous timing. This apparent paradox highlights the importance of integrating objective phase markers with behavioral data, as reliance on reported bedtime alone may obscure the true direction of circadian misalignment. It also raises the possibility that some cases reflect reduced sensitivity to melatonin onset combined with delayed behavioral sleep timing, producing a rebound alertness effect when sleep is attempted well past the optimal biological window, rather than true circadian phase advancement. Alternatively, these profiles may indicate weak or ineffective circadian entrainment. Accordingly, advanced DLMO profiles should be interpreted within a broader symptom, environmental, and behavioral context, as they may signal circadian dysregulation rather than Advanced Sleep Phase Disorder (ASPD) alone.

**Figure 3.**
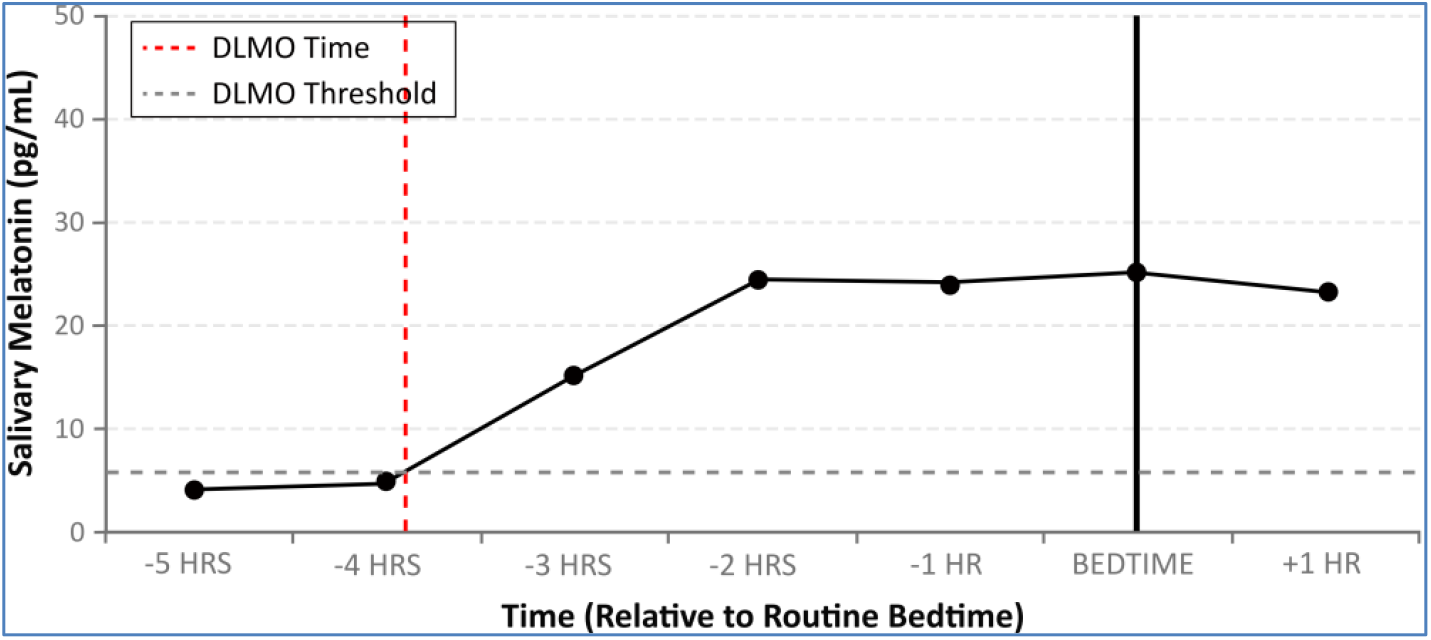
Advanced Melatonin Onset Profile. Representative profile within the Advanced Melatonin Onset group where DLMO is calculated to be > 3 hours prior to bedtime. Vertical dashed line represents DLMO time and horizontal dashed line represents either the calculated or cutoff threshold.

Twenty-seven participants exhibited elevated melatonin levels (>10 pg/mL) across more than 85% of samples, often without a clear physiological onset pattern, even several hours before reported bedtime (see Figure 4). These profiles may reflect severe circadian phase misalignment, in which DLMO occurred prior to the sampling window. However, findings from 24-hour melatonin profiles reinforce that many individuals exhibit sustained elevations across the entire circadian cycle, indicating that these patterns frequently extend beyond missed onset timing alone. Sustained melatonin elevations may result from continued exogenous supplementation, medication effects (e.g., CYP1A2 inhibition or altered serotonin metabolism), impaired hepatic or renal clearance, or physiological alterations in circadian entrainment, receptor sensitivity, or pineal output. The clinical implications of these profiles remain incompletely understood. They may be associated with increased daytime sleepiness or difficulty maintaining alertness, though such symptoms are often attributed to mood disorders or medication side effects. Paradoxically, elevated melatonin levels may coexist with symptoms typically associated with low melatonin output, potentially reflecting reduced circadian amplitude and impaired distinction between sleep and wake states. Hypermelatoninemic profiles may therefore represent an important physiological signal warranting further clinical investigation, particularly given their prevalence in this sample.

**Figure 4.**
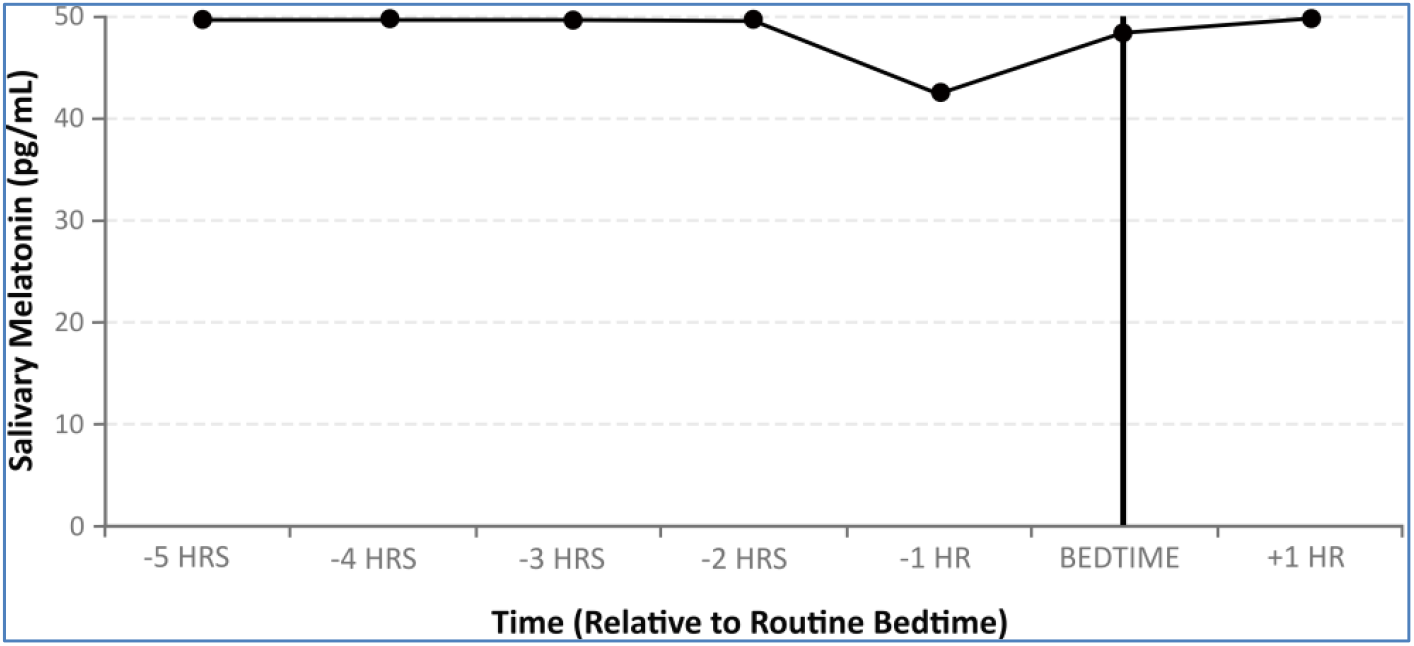
Hypermelatoninemia Profile. Representative profile within the Hypermelatoninemia group where melatonin levels remain elevated above 10 pg/mL for a minimum of 85% of samples; DLMO cannot be reliably calculated.

Thirty-six participants exhibited little to no consistent rise in melatonin, with levels remaining below 10 pg/mL across all samples (see Figure 5). These profiles may reflect medication-related suppression (e.g., β-adrenergic blockers [22]), pineal dysfunction, reduced endogenous production (potentially age-related), or insufficient dim-light control during sampling. Participants exhibiting this phenotype were generally contacted to confirm protocol adherence. Although an extremely advanced or delayed onset occurring outside the sampling window cannot be excluded, the absence of a measurable rise may also indicate compromised circadian signaling. Such deficits could contribute to difficulty initiating or maintaining sleep, poor sleep quality, or impaired circadian entrainment, yet may be misattributed to insomnia, anxiety, or behavioral factors in the absence of objective circadian assessment. Low melatonin profiles may therefore serve as a clinically relevant physiological marker deserving further evaluation.

**Figure 5.**
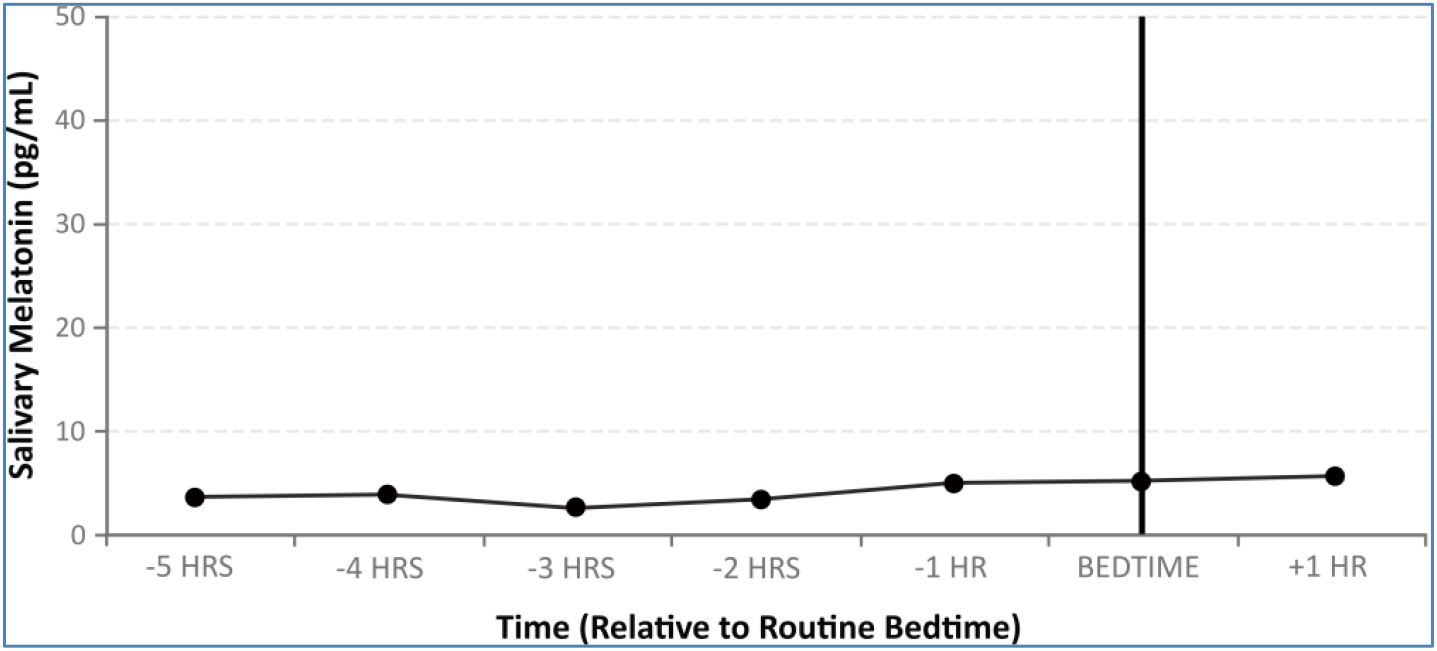
Hypomelatoninemia Profile. Representative profile within the Hypomelatoninemia group where melatonin levels remain below 10 pg/mL for a minimum of 85% of samples; DLMO cannot be reliably calculated.

The five phenotypes described above accounted for 80.5% of cases. However, melatonin profiles in 43 participants appeared unstable, with multiple small peaks or erratic fluctuations (see Figure 6). Two subgroups were evident. First, irregular profiles may arise from methodological artifacts or suboptimal protocol adherence but may also reflect genuine circadian instability. Potential contributing factors included medication timing (93% reported use of medications known to influence melatonin levels, including 72% taking psychiatric medications), recent food intake, and sample handling variability. These findings suggest that, beyond methodological limitations, many individuals may experience real-world challenges in generating or maintaining a stable melatonin signal. Circadian irregularity itself may therefore represent a meaningful clinical feature rather than merely a data collection limitation.

**Figure 6.**
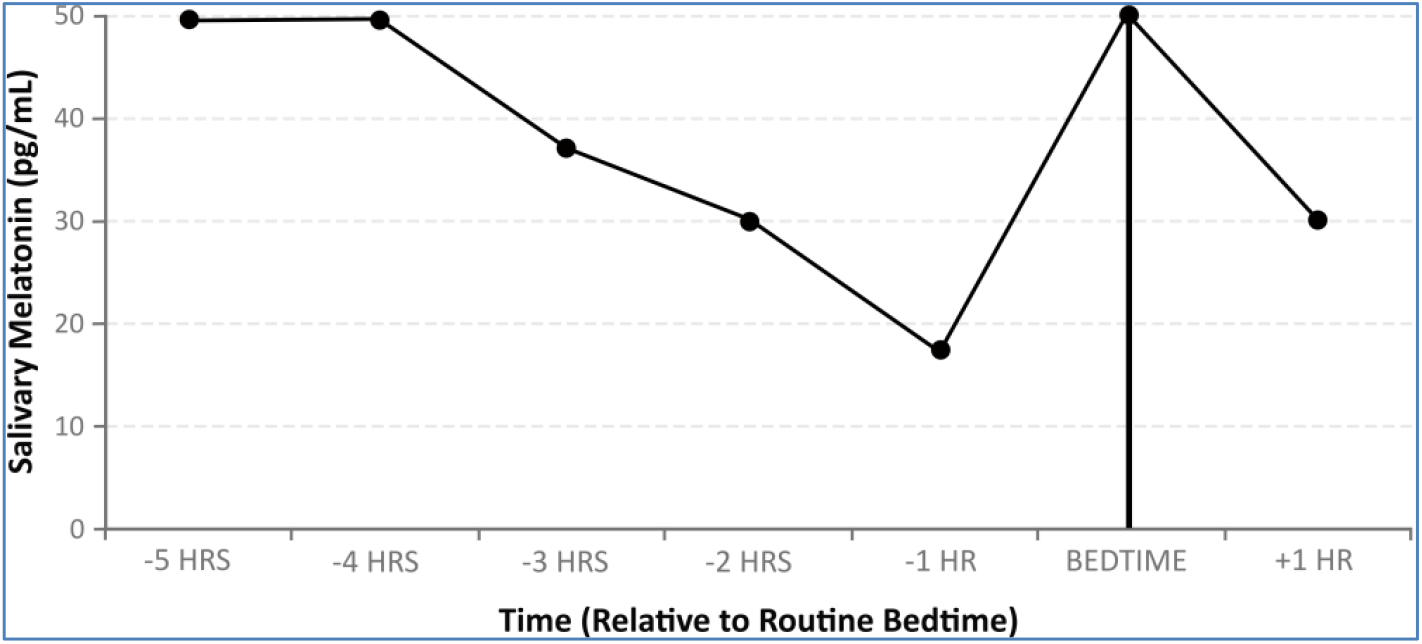
Irregular/Multipeak Profile. Representative profile within the Irregular/Multipeak group, in which melatonin levels remain inconsistent with expected physiological onset patterns; DLMO cannot be reliably calculated.

A second subgroup displayed melatonin profiles characterized by a gradual, steadily increasing trend across the sampling window, without a clear physiological inflection point or distinguishable rise above baseline (see Figure 7). These patterns may reflect onset occurring prior to sampling, elevated or variable baseline melatonin obscuring DLMO detection, or genuinely atypical secretion dynamics. Additional contributors—such as individual variation in melatonin amplitude, environmental masking, or assay sensitivity limitations—cannot be excluded. Given the limited interpretability of these profiles, participants were advised to repeat sampling with an earlier start time and heightened attention to collection conditions.

**Figure 7.**
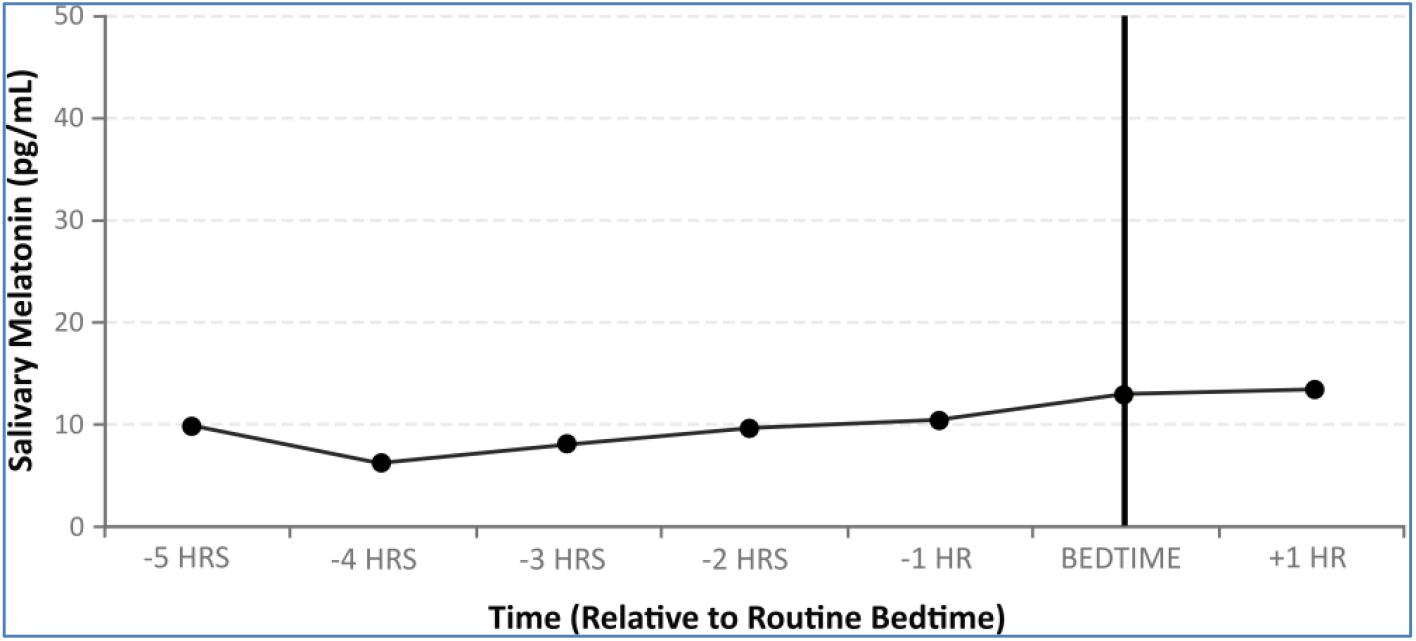
Other/Unclassified Profile. Representative profile within the Other/Unclassified group in which melatonin levels do not exhibit a clear physiological inflection point or a statistically distinguishable rise above baseline; DLMO cannot be reliably calculated.

When stratified by medication use, 78.4% of participants with atypical melatonin profiles reported taking melatonin-modulating medications or supplements, compared with 56% of those exhibiting normative profiles. Psychiatric medications were disproportionately represented among participants with irregular or flat profiles, while β-adrenergic blockers have been shown to suppress melatonin production across the circadian profile [22, 23].

### Limitations

The melatonin profiles studied here were self-collected in in-home environments, which may have introduced variability related to ambient light exposure, food intake, or adherence to sampling timing. The sample consisted of self-and physician-referred individuals with chronic sleep complaints, which may limit generalizability to broader populations without such concerns. The cross-sectional design precludes determination of the stability or reproducibility of observed phenotypes across nights. Time of year and geographic location may influence melatonin secretion through differences in photoperiod and light exposure patterns; however, these factors were not systematically evaluated in this initial sample.

In real-world settings, medication use is ubiquitous. Accordingly, consideration should be given to adjusting medication timing or administering medications outside the sampling window, when safe and feasible, as a best-practice recommendation. The presence of “non-specific” profiles underscores the importance of distinguishing between methodological constraints and true biological variation when interpreting melatonin data. In practice, these potential sources of variation should be carefully considered.

### Conclusion

Taken together, these findings underscore the utility of salivary DLMO profiling for identifying biologically relevant variation in circadian phase and amplitude. This assessment may therefore provide objective physiological context for non-specific sleep–wake complaints that often present with similar symptoms across the sleep–wake cycle. Profiles that deviate from normative patterns may be associated with specific behaviors, comorbidities, or pharmacologic influences. When interpreted within the appropriate clinical context, these patterns may offer a framework for guiding therapeutic strategies, including optimization of sleep timing, behavioral interventions, light therapy, appropriately timed melatonin administration, and scheduling of other pharmacologic treatments to minimize circadian disruption. Finally, efforts to define the predictive value of circadian profiles and to develop clinical guidelines to support their implementation in medical practice represent an important next step.

## Data Availability

All data produced in the present study are available upon reasonable request to the authors

## Acknowledgements

We thank the many physicians, health care providers, caregivers, and patients who participated. We thank Dr. Douglas A. Granger for his editorial contributions.

## REFERENCES

1. Murray, J.M., et al., A Protocol to Determine Circadian Phase by At-Home Salivary Dim Light Melatonin Onset Assessment. J Pineal Res, 2024. 76(5): p. e12994.

2. Abbott, S.M., R.G. Malkani, and P.C. Zee, Circadian disruption and human health: A bidirectional relationship. Eur J Neurosci, 2020. 51(1): p. 567–583.

3. Abbott, S.M. and P.C. Zee, Circadian Rhythms: Implications for Health and Disease. Neurol Clin, 2019. 37(3): p. 601–613.

4. Roenneberg, T., R.G. Foster, and E.B. Klerman, The circadian system, sleep, and the health/disease balance: a conceptual review. J Sleep Res, 2022. 31(4): p. e13621.

5. Lewy, A.J. and R.L. Sack, The dim light melatonin onset as a marker for circadian phase position. Chronobiol Int, 1989. 6(1): p. 93–102.

6. Voultsios, A., D.J. Kennaway, and D. Dawson, Salivary melatonin as a circadian phase marker: validation and comparison to plasma melatonin. J Biol Rhythms, 1997. 12(5): p. 457–66.

7. Pandi-Perumal, S.R., et al., Dim light melatonin onset (DLMO): a tool for the analysis of circadian phase in human sleep and chronobiological disorders. Prog Neuropsychopharmacol Biol Psychiatry, 2007. 31(1): p. 1–11.

8. Keijzer, H., et al., Why the dim light melatonin onset (DLMO) should be measured before treatment of patients with circadian rhythm sleep disorders. Sleep Med Rev, 2014. 18(4): p. 333–9.

9. Gloston, G.F., et al., Integrating Assessment of Circadian Rhythmicity to Improve Treatment Outcomes for Circadian Rhythm Sleep-Wake Disorders: Updates on New Treatments. Curr Sleep Med Rep, 2025. 11(1): p. 8.

10. Bleakley, A., Blunting Occam’s razor: aligning medical education with studies of complexity. J Eval Clin Pract, 2010. 16(4): p. 849–55.

11. McCarty, D.E., Beyond Ockham’s razor: redefining problem-solving in clinical sleep medicine using a “five-finger” approach. J Clin Sleep Med, 2010. 6(3): p. 292–6.

12. Burgess, H.J., et al., Home dim light melatonin onsets with measures of compliance in delayed sleep phase disorder. J Sleep Res, 2016. 25(3): p. 314–7.

13. Burgess, H.J., et al., Home Circadian Phase Assessments with Measures of Compliance Yield Accurate Dim Light Melatonin Onsets. Sleep, 2015. 38(6): p. 889–97.

14. Pullman, R.E., S.E. Roepke, and J.F. Duffy, Laboratory validation of an in-home method for assessing circadian phase using dim light melatonin onset (DLMO). Sleep Med, 2012. 13(6): p. 703–6.

15. Kennaway, D.J., The dim light melatonin onset across ages, methodologies, and sex and its relationship with morningness/eveningness. Sleep, 2023. 46(5).

16. Burgess, H.J. and L.F. Fogg, Individual differences in the amount and timing of salivary melatonin secretion. PLoS One, 2008. 3(8): p. e3055.

17. Lewy, A.J., N.L. Cutler, and R.L. Sack, The endogenous melatonin profile as a marker for circadian phase position. J Biol Rhythms, 1999. 14(3): p. 227–36.

18. Wehr, T.A., et al., Bimodal patterns of human melatonin secretion consistent with a two-oscillator model of regulation. Neurosci Lett, 1995. 194(1-2): p. 105–8.

19. Benloucif, S., et al., Measuring melatonin in humans. J Clin Sleep Med, 2008. 4(1): p. 66–9.

20. Rahman, S.A., et al., Clinical efficacy of dim light melatonin onset testing in diagnosing delayed sleep phase syndrome. Sleep Med, 2009. 10(5): p. 549–55.

21. Lewy, A.J., et al., Melatonin marks circadian phase position and resets the endogenous circadian pacemaker in humans. Ciba Found Symp, 1995. 183: p. 303–17; discussion 317-21.

22. Stoschitzky, K., et al., Influence of beta-blockers on melatonin release. Eur J Clin Pharmacol, 1999. 55(2): p. 111–5.

23. Murphy, D.L., et al., Effects of antidepressants and other psychotropic drugs on melatonin release and pineal gland function. J Neural Transm Suppl, 1986. 21: p. 291–309.

24. Murray, J.M., et al., Prevalence of Circadian Misalignment and Its Association With Depressive Symptoms in Delayed Sleep Phase Disorder. Sleep, 2017. 40(1).

25. Chang, A.M., et al., Sleep timing and circadian phase in delayed sleep phase syndrome. J Biol Rhythms, 2009. 24(4): p. 313–21.

